# Analysis of Data Use Registers published by health data custodians in the UK

**DOI:** 10.1101/2021.05.25.21257785

**Authors:** Nada Karrar, Shahriar Kabir Khan, Sinduja Manohar, Paola Quattroni, David Seymour, Susheel Varma

**Affiliations:** Health Data Research UK

## Abstract

Transparency of how health and social care data is used by researchers is crucial to building public trust. We define “data use registers” as a public record of data an organisation has shared with other individuals or organisations for the purpose of research, innovation and service evaluation, and are used by some data custodians across the United Kingdom to increase transparency of data use. They typically contain information about the type of data being shared, the purpose, date of approval and name of organisation or individual using (or receiving) the data. However, information published lacks standardisation across organisations. Registers do not yet have a consistent approach and are often incomplete, updated infrequently and not accessible to the public. In this paper, we present an empirical analysis of existing data use registers in the UK and investigate accessibility, content, format and frequency of updates across health data organisations. This analysis will inform future recommendations for a data use register standard that will be published by the UK Health Data Research Alliance.

## Introduction

### The UK Health Data Research Alliance

The UK Health Data Research Alliance (the Alliance) is an independent alliance of data providers, custodians and curators dedicated to improving human health by maximising the potential of health data at scale.

Members of the Alliance include National Health Service (NHS) Trusts, charities, research cohorts, biobanks, Health Data Research Hubs^1^ and national data custodians, that have come together to create a UK-wide, federated and co-ordinated approach toward building a health data research infrastructure.

The Alliance vision is to establish best practice for the trustworthy and ethical use of UK health data for research at scale. Convened by Health Data Research UK (HDR UK)^2^, it covers the four nations of the UK and is focused on data sharing for research. It has successfully built alignment across data custodians on Trusted Research Environments^3^, diversity/representativeness in data, public and patient involvement, data and metadata^4^ standards^5^, and streamlined, and harmonised access management processes.

Datasets from Alliance members are discoverable through the Health Data Research Innovation Gateway^6^, with over 600 datasets, covering up to 54.4 million people as at 18th May 2021. Working in partnership with over 22,000 public and patient contributors, the Alliance, Gateway, Health Data Research Hubs and HDR UK Research Programmes are enabling a secure, federated approach to data-enabled health and care breakthroughs including in COVID-19.

### Increasing transparency with Data Use Registers

One of the founding principles of the Alliance is transparency of governance and operations. Publishing a register of projects and/or organisations using the data under a member’s custodianship is a vital demonstration of this principle.

With this work, the Alliance focuses on aligning approaches to improving and standardising the content, format and frequency of publishing data use registers (also referred to as data release registers or list of approved studies). We define a data use register as a public record of data an organisation has shared with other individuals and organisations for the purpose of research, innovation and service evaluation. It typically contains information about the type of data being shared, the purpose, date of approval and name of organisation using (or receiving) the data (for a glossary of key terms, please refer to the supplementary material).

The need for transparency regarding the use of data was highlighted in a recent report ‘Putting Good into Practice’ by the National Data Guardian (NDG) and Understanding Patient Data^7^. The report states that *‘transparency cannot be separated from public benefit’*. As such, the public have a right to know how, why and by whom their data is being used and for this to be possible, ‘*transparency is required throughout the whole data life cycle, not just at the point of application*.’

Therefore, publishing a register of projects, studies and/or requests that an organisation has supported through the provision of data is a demonstration of the requirements outlined by the NDG. Currently, this information is not always made public or there is a significant time lag before such records are published. Moreover, there is a clear lack of consistency when it comes to the content, functionality and purpose of these registers, as also highlighted by custodians, public contributors and researchers during recent workshops hosted on behalf of the Alliance.

In addition to meeting transparency expectations, national bodies have a legal obligation to make this information publicly available. Research databases are also required to share a minimum dataset, as outlined by the ethics committee in their conditions for ethical approval^8^. As such, not only is it important that all data custodians provide access to data for research, it is also crucial that information about this activity is transparent and accessible to the general public, as well as to researchers and funders.

Data use registers also have the potential to improve the efficiency of research by highlighting past and present research projects and data uses to researchers and funders, prioritising funding for national data assets, and identifying under-served areas of data collections or research that could be prioritised for future funding. They could also help close the loop on demonstrating public benefit by linking to the outputs and outcome of data use.

Here we present an empirical analysis of some of the existing data use registers in the UK and investigate accessibility, content, format and frequency of updates across health data organisations. This analysis will inform future recommendations for a data use register standard that will be published by the Alliance.

## Methodology

To inform recommendations for data use registers we reviewed 46 data custodians and controllers from the Alliance and 7 non-Alliance members. While the Alliance was selected as our primary cohort of UK health data custodians that could be used for analysis, seven non-Alliance members were also reviewed to capture the perspective of organisations outside of the Alliance. The analysis of data use registers was performed on a total of 28 registers, 22 from Alliance members and 6 from non-Alliance members. A full list of the data custodians reviewed is provided in Table 1.

**Table 1.**
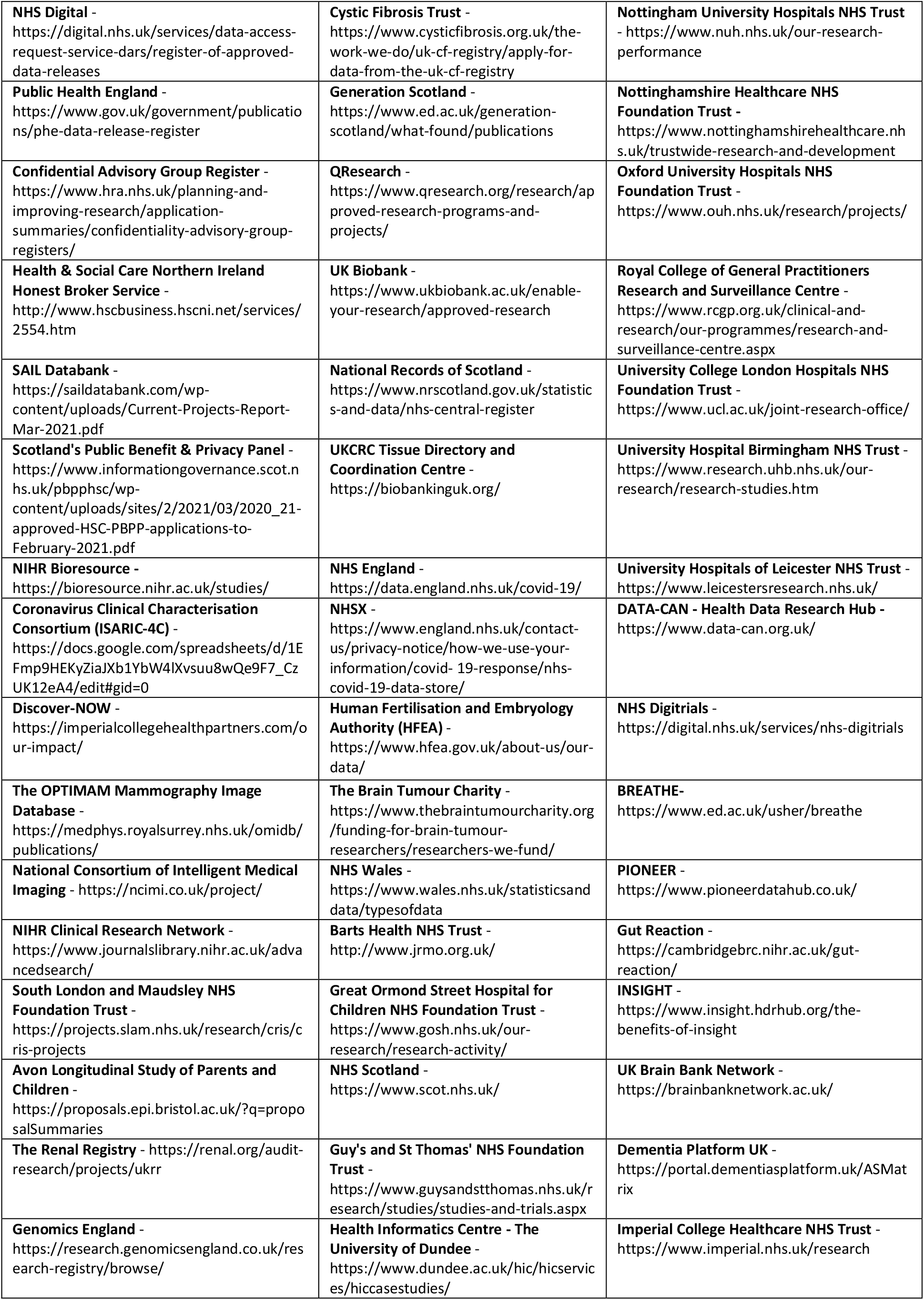
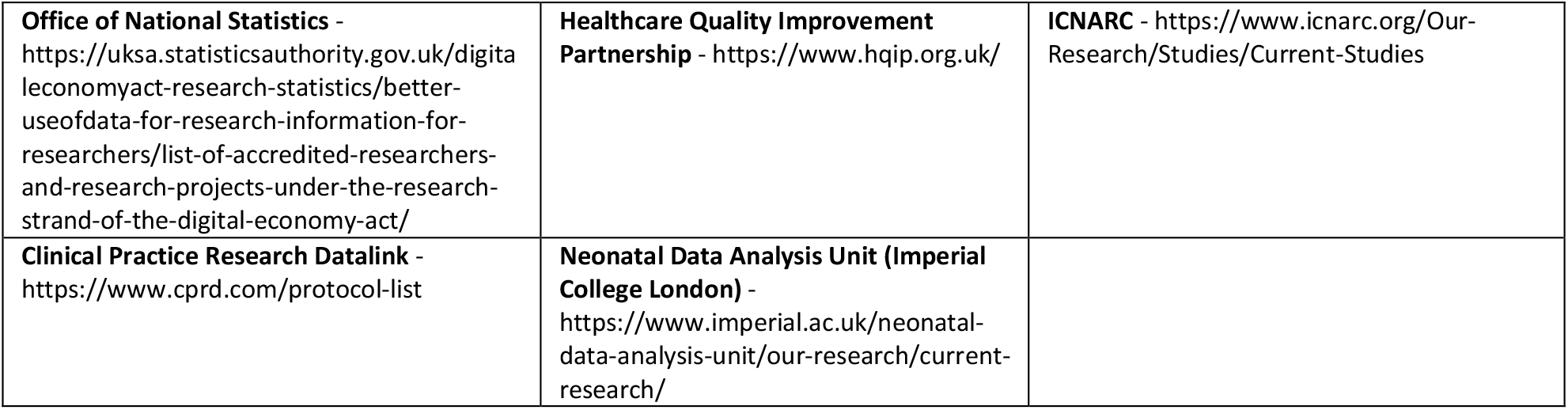
List of 53 data custodians included in the analysis, along with a link to their data use register or homepage if not available.

The aim of this analysis is to understand the current state of transparency and level of information provided on health data usage, with the view to inform future recommendations to be published by the Alliance.

### Data Collection

#### Inclusions

Data custodians and controllers responsible for the safe collection, storage and sharing of health data for research and innovation. The type of organisations included are NHS trusts, medical charities, research cohorts, biobanks, health data research hubs and national data custodians (Table 1).

Of the 53 data custodians included in this review, 46 were Alliance members and 7 were non-Alliance data custodians and controllers or other organisations with a responsibility for decisions about data use (e.g., Health Research Authority). Analysis was performed on 28 data use registers.

#### Exclusions

Clinical trial registers were excluded as the scope of this analysis was limited to registers of approved data access requests, rather than the recruitment of participants to clinical trials.

#### Limitations

The 53 data custodians included in the review is not an exhaustive list of all health data custodians. As we focused mainly on Alliance members there are likely to be data custodians that have been omitted.

In addition, although each data custodian’s website was extensively reviewed for a data use register, there may be a public record of approved data use requests that we were unable to locate.

#### Quantitative data

Secondary data was collected over a period of two weeks (06-May-2021 to 18-May-2021) through a review of data custodian websites. The content of each data use register was checked for the availability of 30 fields. Each field was grouped against the Five Safes Framework^9^ on data access (Safe People, Safe Projects, Safe Data, Safe Settings, Safe Outputs - see Table 2).

**Table 2.** Summary table of the 30 checks made on the content of each data use register, along with a definition of each check and it’s respective Five Safe domain.

| Five Safes | Check to be Made | Definition |
| --- | --- | --- |
| **Safe People** | Organisation Name (LIST) | The name of the legal entity that signs the contract to access the data |
|  | Funders/Sponsors/ Collaborators (LIST) | The name of any funders, sponsors or collaborators involved in the project |
|  | Organisation Type | The type of organisation that has signed a contract to access the data |
|  | Applicant Name (LIST) | The name of the principal applicant that has been authorised to use the data |
|  | Accredited Researcher Status | The accreditation status of a researcher, as defined by the ONS Research Code of Practice and Accreditation criteria (see supplementary material for pre-defined values) |
|  | Organisation ID | A unique identifier for an organisation that is preferably an industry used standard, such as Grid.ac<sup>16</sup> |
|  | Applicant ID | A unique identifier for the applicant that is preferably an industry used standard, such as Grid.ac |
| **Safe Projects** | Project Title | The title of the project/research study/request that the applicant is investigating through the use of health data |
|  | Lay Summary | A concise and clear description of the project, (e.g. as required by UKRI<sup>17</sup> in funding applications). It should outline the problem, objectives and expected outcomes in language that is understandable to the general public |
|  | Public Benefit Statement | A description in plain English of the anticipated outcomes, or impact of project on the general public |
|  | Approval Date | The date the application was approved |
|  | Legal Basis for Provision of Data<sup>18</sup> | The legal basis that allows the applicant to lawfully process personally identifiable data (see supplementary material for pre-defined values) |
|  | Common Law Duty of Confidentiality | In the application of the Common Law Duty of Confidentiality there are 2 options that enable a release: Consent (reasonable expectation) or Section 251 NHS Act 2006<sup>19</sup> |
|  | Release Date | The date the data was released |
|  | Project ID | A unique identifier for the project that is preferably an industry used standard, such as IRAS ID<sup>20</sup>. However, for non-research projects, a unique reference number created by the data custodian on receipt of the application is sufficient |
|  | Technical Summary | A summary of the proposed research, in a manner that is suitable for a specialist reader |
|  | Approval Status | Current stage of the applicant's request for access |
|  | Other Approval Committees (LIST) | Reference to other decision-making bodies that the project has already been authorised by |
|  | Project Start Date | The date the project is scheduled to start or actual start date |
|  | Project End Date | The date the project is scheduled to finish or actual end date |
| **Safe Data** | Dataset(s) Name | The name of the dataset(s) being accessed, as determined by the data controller |
|  | Data Sensitivity Level | The level of identifiability of the data being accessed, as defined by Understanding Patient Data |
|  | Request Category Type | This categorises the 'purpose of the share' (i.e. research, policy development, etc) |
|  | National Data Opt-Out Applied? | Specifies whether the preference for people to opt-out of their confidential patient information being used for secondary use has been applied to the data prior to release |
|  | Request Frequency | Determines whether this a 'one-off' request or a recurring dataset to be provided over a specific time period |
|  | Description of how the data will be used? | Details of how the data requested will be used (including how they will analyse the data or link it to other datasets) |
|  | Description of Confidential Data Used | A description of the specific patient identifiable fields that have been included in the dataset(s) being accessed |
|  | Publisher Name | The name of the organisation responsible for running or supporting the data access request process |
| **Safe Settings** | Trusted Research Environment or any other specified location | These are highly secure spaces for researchers to access sensitive data (also known as Data Safe Havens). If a TRE has not been specified, is their mention of any other location for data usage? |
| **Safe Outputs** | Link to Research Outputs | A link to any academic or non-academic research outputs, as they become available |

We also collected data on register format and publication frequency, as evidenced by the last known update.

The ease of both finding and searching through the register, along with the ability to download or export all of the data was captured.

Finally, as websites and registers are subject to change, we documented the date of each review (along with any additional useful findings in the comments section).

#### Analysis

Statistical analysis was used to calculate the total number and percentage of data custodians within the sample size that met the definition of each check. Please refer to the raw data file for specific calculations (Dataset 1, 10.5281/zenodo.4783490^10^).

## Results

Here we present the results of our analysis of the current state of data use registers published by data custodian organisations.

### What proportion of Alliance members have a public data use register?

Of the 46 Alliance members analysed, 48% (22) had data use registers that were discoverable via public websites (Figure 1).

**Figure 1.**
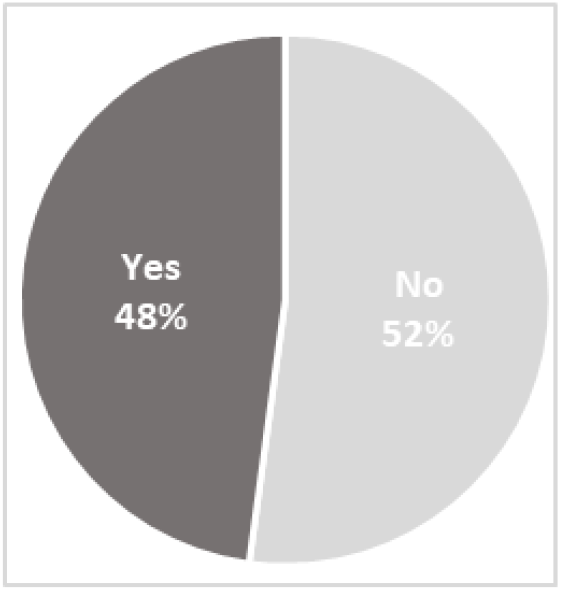
Percentage of Alliance members currently publishing data use registers. Of the 46 Alliance members analysed, 22 (48%) had data use registers that were discoverable via a public website, based on our internet search.

### What information is included in the data use register?

Figure 2 shows the information we found to be most commonly included in the 28 data use registers identified.

**Figure 2.**
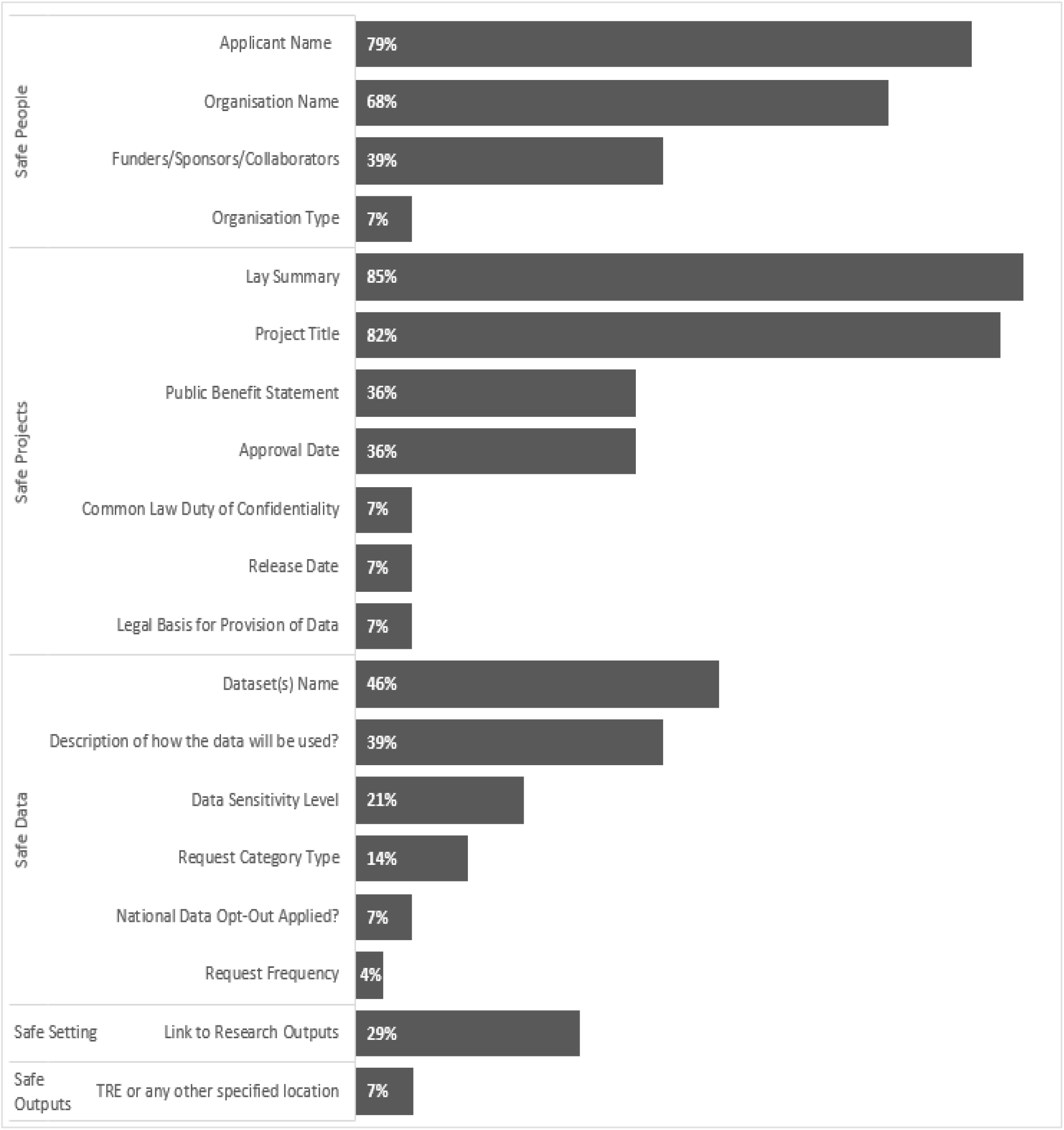
Information most commonly included in the 28 data use registers identified. The percentage of data custodians that have included the 19 most common fields in their data use register, grouped according to the Five Safes Framework specified in Table 2.

Lay summary was the most common feature in all the registers analysed (85%). However, it is important to highlight that many of these summaries were not in plain English and did contain technical or scientific language. Assessing the quality of lay summaries was not within the scope of this analysis, the inclusion of a non-technical summary was deemed to meet the requirements of a lay summary.

Details on the applicant and their associated organisation were also very common, 79% and 68% respectively. By contrast, only a third of data custodians included information on the public benefits of the approved data use requests. Similar to information on funders, sponsors or collaborators (39%).

Although nearly half (46%) of data custodians made reference to the dataset(s) being shared or accessed, there was less transparency on the sensitivity level of this data (21%).

Information on the legal basis for data sharing, as well as the application of the Common Law Duty of Confidentiality^11^ and national data opt-outs^12^ was the least common, with only two data custodians providing this data. Whilst this is a core requirement for national data custodians, it is worth noting that in the case of anonymised data, not all data uses will require legal justification. Moreover, the opt out may have been applied ‘upstream’ in the data processing pathway.

### What format are the data user registers in?

Our web-based analysis showed that 68% (19) of data use registers were published on a web page in either a list or table format; 18% (5) were found as a link to a spreadsheet available for download; 14% (4) were found as a link to a pdf available for download (Figure 3).

**Figure 3.**
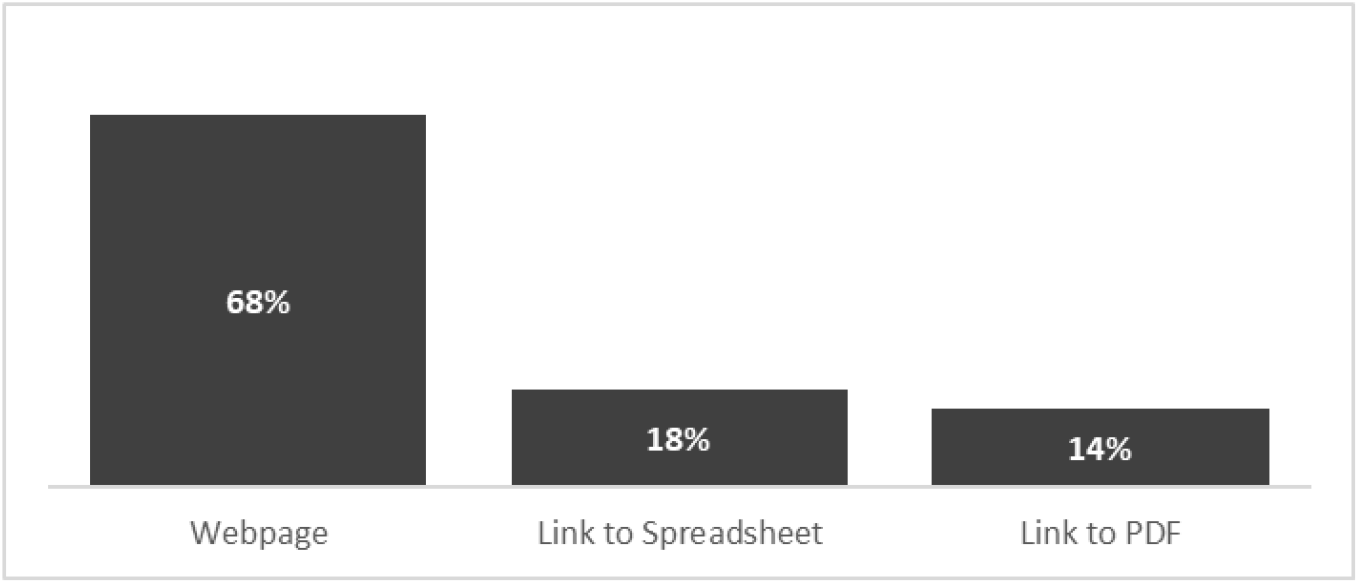
Data use register format most commonly found. Of the 28 data use registers located, 19 (68%) were published directly on the webpage via a table or list, 5 (18%) were linked to a spreadsheet and 4 (14%) were linked to PDF.

### How frequently are the data use registers updated?

We found that information on the frequency of publication was not directly communicated by the majority of data custodians, with 68% (19) not specifying how often their registers were updated. Of the nine data use registers that did share this information or provide a last known entry date, three are updated daily, one weekly, three monthly and two quarterly (Figure 4).

**Figure 4.**
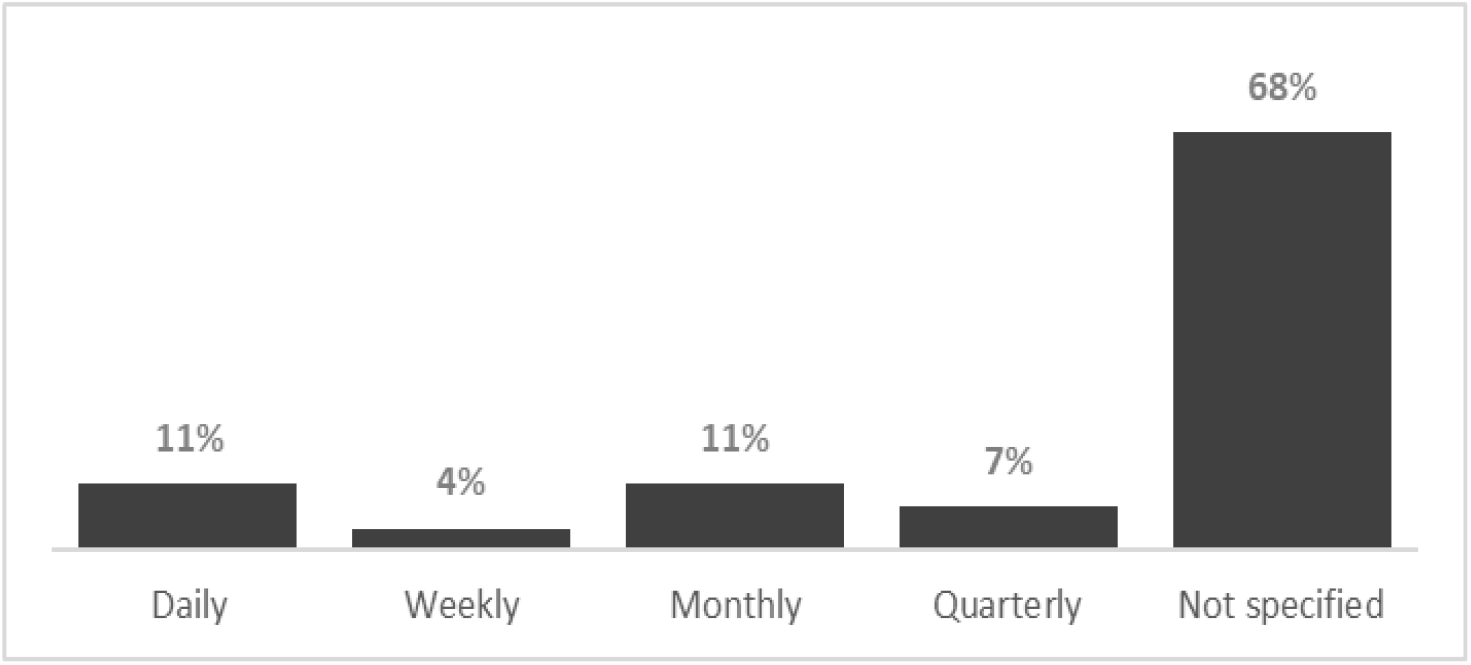
Frequency of updates to data use registers. Of the 28 data use registers identified, 18 (68%) did not specify how often the register is updated or offer a last known entry date. Three (11%) were updated daily, 1 (4%) weekly, 3 (11%) monthly and 2 (7%) quarterly.

### How findable, searchable and downloadable are the registers?

The majority of data use registers (71%) were relatively easy to locate. An easily locatable register was defined by having an intuitive and quick user journey from the homepage to the data use register. However, slightly fewer registers (43%) provided an in-built search function. Less than a third of the data use registers reviewed allowed all of their content to be downloaded (Figure 5).

**Figure 5.**
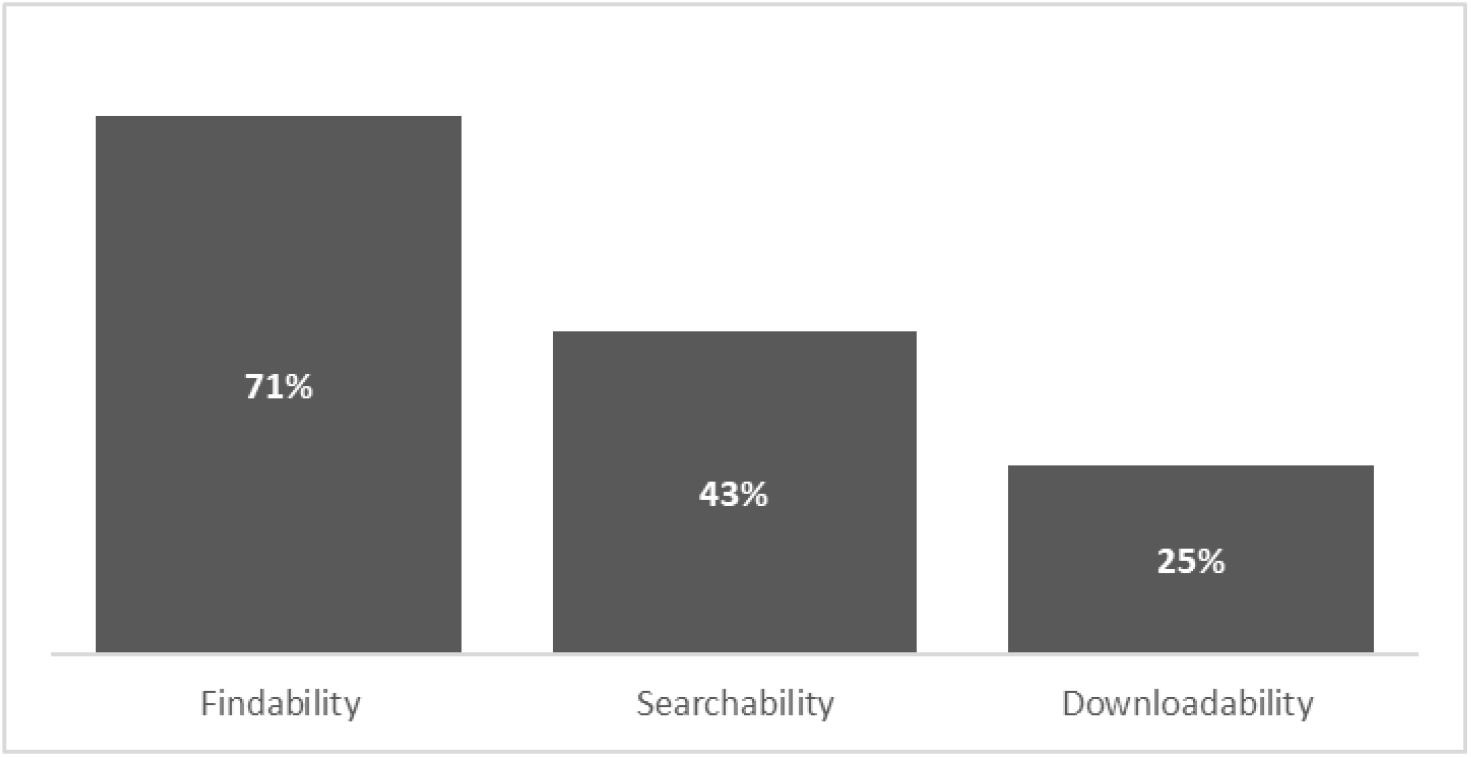
Findability, searchability and downloadability of data use registers. ‘Findability’ is defined as a register that is quick and easy to locate, of which 20 (71%) met this definition; ‘searchability’ is defined as having a search function available within the register, which 12 (43%) registers provided; ‘downloadability’ is defined as having all data within the register available for download, of which 7 (25%) data use registers allowed.

## Discussion

Public trust in the way data is shared for research and for the purpose of running public services is vital to ensure data driven technologies can accelerate progress and deliver public benefit. Adequate safeguards and governance arrangements are currently in place for collection and use of personal information that enables sharing of data without undermining individual privacy and patient confidentiality^13^. To build public trust and demonstrate trustworthiness, information about how data is accessed and used, as well as the purpose of using this data needs to be effectively communicated - however it is not always accessible and transparent.

An independent report published by the Centre for Data Ethics and Innovation^14^ highlighted that the benefits of data sharing are not always felt equally among people, with some thinking that data is not used for the explicit and direct benefit of individual citizens. To reassure the public and increase trust, transparency of processes and clarity around purpose and impact of data uses is fundamental. Data use registers can help build and improve public knowledge and understanding while demonstrating the purpose of using confidential information for public benefit and the impact that might have been derived from it.

Our research on the current state of data use registers has provided valuable insight into the practices of data controllers and custodians with established data use registers. While half of the custodians analysed were found to publish registers, the others do not. This was of biggest concern to the public contributors involved who then highlighted in discussions that the first important implementation step will be to support organisations without a register to make this information public and overcome any blockers or barriers.

There is a lot of variability in the content, format and frequency of publication, which demonstrates the need for a consistent approach through a standard and provides major scope for improvement.

There is also an opportunity for behavioural changes amongst data custodians and researchers. Introducing the link to research outputs supported by the ability of assigning Digital Object Identifiers to datasets (as shown by 29% of the data use registers reviewed, Figure 2) has the potential to showcase the impact and benefit of data used for research. Demonstrating clear links between data use and research impact should help build the confidence of patients and the public in sharing their data and create a culture of transparency and openness. With this work we hope to inform best practice and highlight the relevance of data citation in data management practice, where alignment between researchers, data custodians and funders becomes necessary.

An added layer of depth and context was also gained through the knowledge and experience shared by our stakeholders during workshops and via surveys. Members from our patient and public involvement networks, highlighted that data accuracy and authenticity is essential to building their confidence and trust. It was suggested that automated processes could facilitate the transfer of information from data access management systems to data use register, making it easier to keep data use registers up to date and ensure accurate information is publicly shared.

The feedback received to date by researchers, data custodians and the public has confirmed the need and value of data use registers in improving transparency in the use of health data for research, building public trust, as well as highlighting priorities and challenges likely to be faced in implementation. The analysis of current data use registers also provided an overview of elements that could be improved.

The activities completed in this discovery phase of the project have been key to shaping and informing an emerging set of recommendations on the data use registers standard that will be published on behalf of the Alliance. These recommendations will be shared with the wider community through the publication of a green paper for consultation. In addition, further engagement with key stakeholders within and outside of the Alliance will help share learning and best practices, as we strive to develop a minimum standard that can be adopted by data custodians nationwide.

## Data Availability

The data underlying this paper are available in Zenodo: https://doi.org/10.5281/zenodo.4783490 Data are available under the terms of the Creative Commons Zero "No rights reserved" data waiver.

## Author contribution

P.Q. and D.S. conceptualised the idea and overarching goals and aims. N.K. developed the theory, coordinated the research activities, performed analysis and wrote the paper. K.K. was responsible for conducting the online research and data collection. S.V. validated the research activities and outputs. P.Q. provided oversight and leadership responsibility for this work, and reviewed the paper. S.M. contributed to the writing and reviewed the paper. All authors discussed the results and contributed to the final manuscript.

## Acknowledgements

We thank all organisations involved in discussions around standards for data use registers.

## Data availability

The data underlying this paper are available in Zenodo: https://doi.org/10.5281/zenodo.4783490 Data are available under the terms of the Creative Commons Zero “No rights reserved” data waiver.^15^

