## Supplementary Material for "Analysis of Data Use Registers published by health data custodians in the UK"

### Glossary of key terms

| Field | Definition |
|---|---|
| **Accredited Researcher Status**[i] | The accreditation status of a researcher, as defined by the ONS Research Code of Practice and Accreditation criteria. |
| **Full Accredited Researcher**[ii] | To be a full accredited researcher, individuals must have an undergraduate degree (or higher) including a significant proportion of maths or statistics or be able to demonstrate at least three years quantitative research experience.<br>Successful completion of a Safe Researcher training course is also required as part of the accreditation process. Full accredited researcher status is valid for five years. |
| **Provisional Accredited Researcher**[iii] | A provisional accredited researcher under the Digital Economy Act (DEA) can work on projects in the Secure Research Service (SRS) under the supervision of an accredited researcher. This role suits individuals without the appropriate experience or qualifications required for full accredited researcher status.<br>Successful completion of a Safe Researcher training course is still required. Provisional accredited researcher status is now valid for three years |
| **Non-Accredited Researcher** | Does not meet the full or provisional accreditation status, as defined by the DEA. |
| **Applicant ID** | A unique identifier for the applicant that is preferably an industry used standard such as Grid.ac[iv] . |
| **Applicant Name** | The name of the principal applicant that has been authorised to use the data (in format First Name, Last Name). |
| **Approval Date** | The date the application was approved in ISO 8601 format. |
| **Data (Patient)**[v] | Data that is collected about a patient whenever they go to a doctor or receive social care. It may include details about the individual's physical or mental health, such as height and weight or detail of any allergies, and their social care needs and services received. It may also include next of kin information. This is recorded and stored in a care record. |
| **Data Controller**[vi] | A term used to describe an individual or organisation who determines the purposes for which and the manner in which any personally identifiable data is or will be processed. It is the responsibility of the Controller to ensure that any processing of personally identifiable data is lawful. |
| **Data Custodian** | Data Custodians are responsible for the safe custody, transport, storage of, and access to data. |
| **Data Processor** [vii] | A term used to describe any person or organisation (other than an employee of the controller) who processes personally identifiable data on behalf of the controller. Controllers must choose to have a written contract (detailing the information governance requirements) in place and have effective means on monitoring, reviewing and auditing their processing. |
| **Data Sensitivity Level** | The level of identifiability of the data being accessed. |
| **Personally Identifiable** [viii] | This term describes personal information about identified or identifiable individuals, which should be kept private or secret. It includes the definition of personal data in the Data Protection Act, but also includes data relating to people who have died and information given in confidence under the Duty of Confidentiality. Identifiers include: name, address, full postcode, date of birth or NHS number. |
| **De-Personalised** [ix] | This is information that does not identify an individual, because identifiers have been removed or encrypted. However, the information is still about an individual person and so needs to be handled with care. It might, in theory, be possible to re-identify the individual if the data was not adequately protected, for example if it was combined with different sources of information. |
| **Anonymous** [x] | This is information from many people combined together, so that it would not be possible to identify an individual from the data. It may be presented as general trends or statistics. Information about small groups or people with rare conditions could potentially allow someone to be identified and so would not be considered anonymous. |

| | |
|---|---|
| **Data Use Register** | A public record of studies, projects and requests that a data custodian or controller has supported through the provision or access of health data for research, innovation and service evaluation. |
| **Dataset(s) Name** | The name of the dataset(s) being accessed, as determined by the data controller. |
| **Description of Confidential Data Used** | A description of the specific patient identifiable fields that have been included in the dataset(s) being accessed. |
| **Funders/Sponsors/Collaborators** | The name of any funders, sponsors or collaborators involved in the project. |
| **Lay Summary** | A concise and clear description of the project, (e.g. as required by UK Research and Innovation[xi] in funding applications). It should outline the problem, objectives and expected outcomes in language that is understandable to the general public |
| **Legal Basis for Provision of Data** [xii] | The legal basis that allows the applicant to lawfully process personally identifiable data, as specified by NHS Digital. |
| **Health and Social Care Act 2012 - s261(2)(c)**[xiii] | Consent of the individuals involved in research is the basis for release of data. |
| **Health and Social Care Act 2012 - s261(7)** | Release of data on the basis of an exemption from the Common Law Duty of Confidence through use of section 251 of the National Health Service Act 2006 and its current Regulations, the Health Service (Control of Patient Information) Regulations 2002. |
| **Health and Social Care Act 2012 - s261(1) and s261(2)(b)(ii)** | Public interest is appropriate for data to be released in pseudonymised format. |
| **Health and Social Care Act 2012 - s261(5)(d)** | NHS Digital releases data to organisations for the purpose of exercising functions conferred by legislation. |
| **Health and Social Care Act 2012 - s261(4)** | NHS Digital releases data to organisations that could have received the data in the first place. |
| **Health and Social Care Act 2012 - s261 - Other dissemination** | Public interest is appropriate for data to be released in pseudonymised format. |
| **Approved researcher accreditation under section 39(4)(i) and 39(5) of the Statistical Registration Service Act 2007**[xiv] | NHS Digital is required to release data as part of the Office for National Statistics (ONS) Approved Researcher Scheme. To access data in this way, an individual must hold ONS Researcher Accreditation and have their research proposal approved by the ONS Microdata Release Panel, on behalf of the National Statistician. |
| **Section 36, Health Act 2009**[xv] | NHS Digital is required to release data in support of collection of tax for NHS staff and contractors. |
| **Section 42(4) of the Statistics and Registration Service Act (2007) as amended by section 287 of the Health and Social Care Act (2012)**[xvi] | NHS Digital is required to release data under the Statistics and Registration Service Act 2007 in support of management of registers of birth and death. |
| **Linkage**[xvii] | The merging of information or data from two or more sources, with the object of combining facts concerning an individual or an event, which are not available in any separate record. |
| **Linked Dataset** | This specifies whether the dataset being accessed is comprised of multiple linked data assets |
| **National Data Opt-Out Applied** [xviii] | A new national data opt-out was introduced in May 2018, following recommendations from the National Data Guardian. People can opt out of having their confidential patient information shared for reasons beyond their individual care, for example for research and planning. |
| **Yes**[xix] | Specifies that 'Yes' an opt-out decision has been upheld and applied to the data prior to release. |
| **Not Applicable**[xx] | The opt-out has not been applied to the data prior to release, as it not applicable for a number of reasons (specified by NHS Digital). |
| **Organisation ID** | A unique identifier for an organisation that is preferably an industry used standard such as Grid.ac[xxi]. |
| **Organisation Name** | The name of the legal entity that signs the contract to access the data. |
| **Organisation Type** | The type of organisation that has signed a contract to access the data |
| **Other Approval Committees** | Reference to other decision-making bodies that the project has already been authorised by. |
| **Project End Date** | The date the project is scheduled to finish or actual end date in ISO 8601 format. |

| | |
|---|---|
| **Project ID** | A unique identifier for the project that is preferably an industry used standard, such as IRAS[xxii]. However, for non-research projects, a unique reference number created by the data custodian on receipt of the application is sufficient. |
| **Project Start Date** | The date the project is scheduled to start or actual start date in ISO 8601 format. |
| **Project Title** | The title of the project/research study/request that the applicant is investigating through the use of health data. |
| **Public Benefit Statement** | A description in plain English of the anticipated outcomes, or impact of project on the general public. |
| **Publisher Name** | The name of the organisation responsible for running or supporting the data access request process. |
| **Release Date** | The date the data was released in ISO 8601 format. |
| **Request Category Type** | This categorises the 'purpose of the share' (i.e. research, policy development, etc.). |
| **Request Frequency** | Determines whether this a 'one-off' request or a recurring dataset to be provided over a specific time period. |
| **Research Outputs** | Any academic or non-academic outputs of the research project. |
| **Technical Summary** | A summary of the proposed research, in a manner that is suitable for a specialist reader. |
| **Trusted Research Environment (TRE)** | These are highly secure spaces for researchers to access sensitive data (also known as Data Safe Havens). |
